# Automating ACMG Variant Classifications Using BIAS-2015: An Algorithm Overview and Benchmark Against the FDA-Approved eRepo Dataset

**DOI:** 10.1101/2024.09.18.24313916

**Authors:** Chris Eisenhart, Joel Mewton, Rachel Brickey, Vafa Bayat

**Affiliations:** Bitscopic Inc., Los Angeles, 90024, CA, USA

## Abstract

In 2015, the American College of Medical Genetics and Genomics (ACMG) in collaboration with the Association of Molecular Pathologists (AMP) published guidelines for the interpretation and classification of germline genomic variants. The ACMG terminology guidelines outlined criteria for assigning one of five categories: benign, likely benign, uncertain significance, likely pathogenic and pathogenic. While the paper laid out 28 different classifiers and the justification for them, it did not provide specific algorithms for implementing these classifiers in an automated manner. Here we present the Bitscopic Interpreting ACMG Standards 2015 (BIAS-2015) software as a complete, open-source algorithm which categorizes variants according to the ACMG classification system. BIAS-2015 evaluates 18 of the 28 ACMG criteria to classify variants in an automated and consistent way while recording the rationale for each classifier to enable in-depth review. We used the genomic data from the ClinGen Evidence Repository (eRepo v1.0.29), one of two FDA-recognized human genetic variant databases, to evaluate the performance of the BIAS-2015 algorithm. All code for BIAS-2015 has been made available on GitHub.

## Introduction

The clinical interpretation of genetic variants is crucial for translating genomic data into actionable clinical practice. In 2015, the American College of Medical Genetics and Genomics (ACMG) and the Association for Molecular Pathologists (AMP) established comprehensive guidelines for the interpretation and classification of germline genomic variants into five categories: pathogenic (P), likely pathogenic (LP), uncertain significance (VUS), likely benign (LB), and benign (B) (Richards et al., 2015). These guidelines are based on 28 criteria grouped into different levels of evidence, such as population data, computational predictions, functional data, segregation data, and *de novo* data. In the years since, these guidelines have become the community standard due to their systematic approach (Amendola et al., 2016).

Despite these guidelines, the interpretation of genetic variants remains a complex and labor-intensive process. Since 2015, many tools have been created to automate this process including InterVar, Sherloc, PathoMAN and others (Li and Wang, 2017; Nykamp et al., 2017; Ravichandran et al., 2019). When deciding to create BIAS we evaluated several factors. We looked for open-source code bases, algorithms that function on freely accessible data and software, algorithms that adhere to the ACMG classifiers and combining criteria, and algorithms that report detailed explanations along with each classifier code. In our analysis of the available software, we did not find any that met all our criteria and so we set out to create BIAS-2015.

BIAS-2015 is built on top of Illumina’s Illumina Connected Annotations (ICA) software (Illumina, 2024). It runs on the ICA JSON (JavaScript Object Notation) output. ICA is a freely accessible CLI annotation software developed by Illumina (previously known as NIRVANA) (Stromberg et al., 2017). Currently, Illumina has a significant market share of all sequencers used in high-end bioinformatics clinical pipelines and many of these labs also run Illumina’s DRAGEN secondary analysis pipeline (Fortune Business Insights, 2024). DRAGEN includes ICA, and therefore these labs will already have the ICA output files for every sequencing run (Behera et al., 2024). Therefore, we decided to build BIAS-2015 using the ICA annotations as a starting point.

The algorithm operates in three phases: the preprocessing step which gathers data from community-standard sources and extracts the information needed for classification, the data loading step that loads in the preprocessing-generated datasets, and the classification phase where the ACMG codes are applied and ACMG combining criteria used to classify each variant.

BIAS-2015 supports user-provided classifiers that can be passed into the algorithm with an optional external call file. This optional external call file enables users to provide values for the 10 ACMG classifiers that BIAS-2015 does not evaluate, to update weights for each classifier, and/or to overwrite BIAS-2015 classifiers.

We evaluated the performance of BIAS-2015 by converting the expert-curated data from the ClinGen eRepo into VCF format and then processed the file using BIAS-2015 (Bioinformatics Research Laboratory, 2024). When comparing the BIAS-2015 classifications of these variants to the classifications reported in eRepo, BIAS-2015 achieved a pathogenic sensitivity of 92.2% and a benign specificity of 96.2%. We show that BIAS-2015 is capable of rapidly classifying annotated variants at an average speed of *>*2,000 variants per second by classifying ∼3,000,000 ClinVar variants (Landrum et al., 2018).

## Methods

### Algorithm Overview

BIAS-2015 takes as input an ICA output JSON file and outputs a TSV file with one row per variant, providing the ACMG classification and detailed rationales for every code that was applied.

BIAS-2015 operates in three distinct phases, ensuring both accuracy and efficiency in variant classification (Fig. 1):

**Fig. 1.**
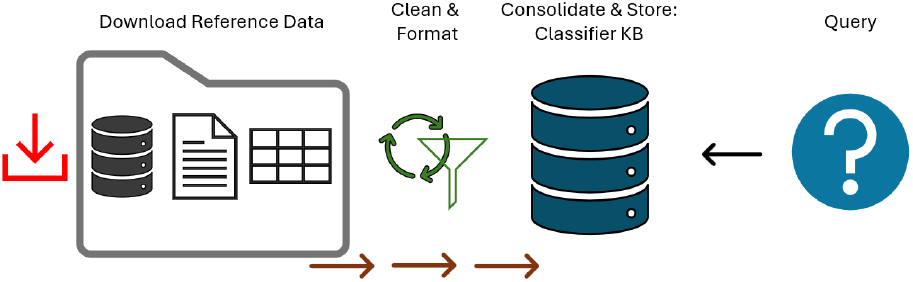
Data Processing Workflow. The process begins with downloading reference data, followed by cleaning and formatting. The refined data is then consolidated and stored in the classifier knowledge base, ready to be queried.

#### 1. Preprocessing Phase

This initial phase involves gathering and compiling data from various external databases, such as the UCSC Genome Browser FTP site (Kent, 2002).

#### 2. Data Loading Phase

When a user runs BIAS-2015 it begins by loading the data files generated via preprocessing. The data from these files is stored in optimized Python data structures, facilitating rapid access during classification.

#### 3. Classification Phase

In this phase, BIAS-2015 iterates over each variant in the JSON file, applying the following 18 ACMG criteria: PVS1, PS1, PS3, PS4, PM1, PM2, PM4, PM5, PP2, PP3, PP5, BA1, BS1, BP1, BP3, BP4, BP6 and BP7 to each variant. The variant is then classified using the ACMG combining criteria.

### ICA Data Extraction

BIAS-2015 leverages the ICA annotations throughout the classification process (Stromberg et al., 2017). This is done by extracting the necessary information from the ICA JSON into a Python data structure that represents a variant, then using this information during classification. BIAS-2015 extracts and uses the following information for each variant: chromosome, position, reference allele, alternate allele, variant type, amino acid change, gene name, consequence, top ClinVar entry, PubMed ID list, gene associated disease, dbSNP ID list, canonical transcript, full transcript list, gnomAD, 1000 Genomes Project, protein domain, GERP, DANN and REVEL scores (Landrum et al., 2018; Sherry et al., 1999; Karczewski et al., 2020; Genomes Project et al., 2015; Davydov et al., 2010; Quang et al., 2015; Ioannidis et al., 2016). Please see the ICA documentation for a detailed explanation of how they define and apply these annotations to each variant.

We use the ICA ClinVar annotations to rank all ClinVar entries with a variant based on classification, submitters, supporting PubMed IDs, expert panel review, phenotypes, and update date. The highest-scoring ClinVar entry is considered for PP5. We use the ICA RefSeq canonical transcript for all classifications.

### Preprocessing and Data File Generation

The ACMG standards and broader clinical bioinformatics community have emphasized the need for tools that can be updated and re-run as new genomic discoveries are made (Richards et al., 2015; Biesecker et al., 2018). To address this need, the BIAS-2015 data files are generated by script using a well-documented and repeatable process. This enables users to update the data files used in classification quickly and in a consistent manner as new updates are released.

The data file generation for each classifier was designed to be expandable to include more lines of evidence as they become available. For instance, though many scripts run off the ClinVar VCF, a user can update the VCF to include their own entries and then re-run the scripts to update the data files. This enables users to incorporate evidence from their own laboratories or projects that may not be included in the ClinVar VCF.

10 of the 18 classifiers BIAS-2015 evaluates (PVS1, PS1, PS3, PS4, PM1, PM4, PM5, PP2, BP1, and BP3) require some form of data preprocessing to support the queries for each of the classifiers.

### ClinVar Review Evidence Weights

BIAS-2015 extensively uses ClinVar during the preprocessing phase to generate data files which are used during classification (Landrum et al., 2018). When evaluating ClinVar variants, BIAS-2015 weights the variants according to their ClinVar review status, where those variants with review status of ‘Practice Guidelines’ and ‘Reviewed by Expert Panel’ are weighted more heavily than variants with review status of ‘Criteria provided, conflicting interpretations’. Variants with no assertion criteria provided are weighted zero and are not included in the final data file. All data files made from ClinVar contain the weights associated with each record, and these weights are then used during classification.

### Classifier Scoring and Weights

The ACMG standards and subsequent SVI publications emphasize weighting classifiers based on their supporting evidence (Richards et al., 2015). They propose using qualifiers such as supporting, moderate, and strong (ClinGen, 2017). To implement this, we have applied a scoring system to each classifier: the classifiers with only the minimum supporting evidence are assigned a score of one (supporting), classifiers with moderate evidence are assigned a score of two (moderate), and classifiers with an abundance of evidence are assigned a score of three (strong). This enables using the combining criteria for pathogenic, likely pathogenic, likely benign and benign laid out by ACMG while incorporating weighting for classifiers with more evidence.

### Pathogenic Classifiers

In the following sections, we describe how BIAS-2015 applies the ACMG criteria to each pathogenic classifier. Figure 2 summarizes which pathogenic classifiers are covered by BIAS-2015 and which should be handled using the optional external call file.

**Fig. 2.**
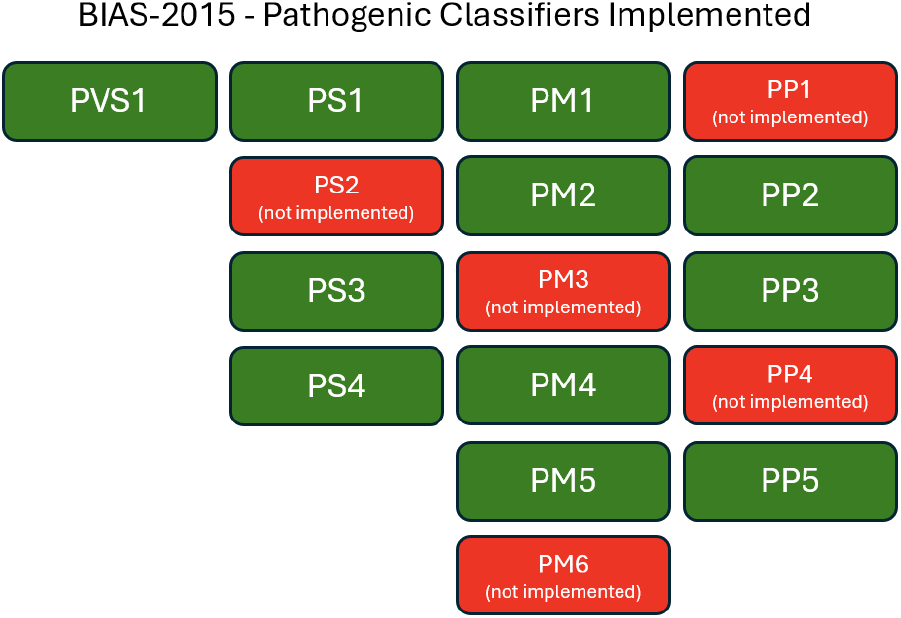
BIAS-2015 Pathogenic Classifiers Implemented. The diagram shows the implementation status of various pathogenic classifiers. Green boxes represent implemented classifiers, while red boxes with “not implemented” indicate classifiers that are not implemented.

#### PVS1

The very strong pathogenic classifier identifies null variants expected to cause significant alteration to the resulting protein in a gene where loss of function (LOF) is a known mechanism of pathogenicity. We began by identifying all genes in gnomAD that have a probability of being LOF intolerant (pLI) greater than 0.8.

To evaluate a variant for PVS1, the variant gene name is checked against the list of LOF genes, and the variant consequence is checked to see if it is known to have a null consequence. If both conditions are met, then PVS1 is applied.

BIAS-2015 evaluates three of five caveats defined by ACMG. The first caveat is handled by our requirement that variants be in our defined LOF gene list. For the second caveat we use the UCSC ncbiRefSeqHgmd.txt file to define the final 50 base pairs in the last coding exon for each gene (Kent, 2002; Pruitt et al., 2005, 2009). This is done in our preprocessing and the resulting data file is loaded at runtime into a mapping table which maps genes to the final 50 bases. To evaluate, we use a variant’s gene name (annotated by ICA) to identify the 50 base region which should be excluded, then check if the variant’s position overlaps the region. If it does overlap, the variant is excluded by caveat two. We do not currently evaluate the third caveat; however, we have plans to integrate splice predictive algorithms such as MMSplice or SpliceAI to enable this (Cheng et al., 2019; Ha et al., 2021; Jaganathan et al., 2019). If there are multiple transcripts, we verify that the consequence seen in the canonical RefSeq transcript is seen in at least one other transcript. If it is not, the fourth ACMG caveat applies. We do not currently evaluate caveat five.

The authors are aware of the SVI PVS1 flowchart and have future aspirations of implementing the logic here as appropriate (Abou Tayoun et al., 2018).

#### PS1

The first strong pathogenic classifier leverages the fact that in most cases when one missense variant is known to be pathogenic, a different nucleotide change that results in the same amino acid change can also be assumed to be pathogenic. We began by identifying the P/LP variants seen in ClinVar without conflicting review status, and then processed these variants with ICA to generate annotation data for each variant (Bhat et al., 2023). The gene and the amino acid change associated with each variant were extracted from this annotated data. This was used to generate a list of P/LP amino acid changes for every gene.

To classify a variant, we check if the gene and amino acid change is seen in our list of pathogenic gene/amino acid relations. If there is a match, the base variant is reported along with the review status of the variant as reported in ClinVar. The score can be reported as a moderate or strong PS1 through two avenues which both represent increased evidence: if the variant was reported as “pathogenic” and/or if there is sufficient weight associated with the ClinVar record.

#### PS3

This classifier looks to bring in variant classification data identified in functional studies, which requires reviewing relevant publications (Brnich et al., 2019). While traditionally a manual step, the recent emergence of artificial intelligence in bioinformatics, specifically in using natural language processing (NLP) algorithms to mine genomic data from publications, has unlocked a treasure trove of information that was previously inaccessible on a widespread, automated scale. The authors recognize that multiple such projects exist, including commercial services such as Genomenon Mastermind (Chunn et al., 2020). For the implementation of this algorithm, the authors used the UCSC AVADA (Automatic Variant Evidence Database) data track: an open-source, academic collaboration that used NLP to identify pathogenic variants from published literature (Birgmeier et al., 2020).

When the data is loaded, all publications are associated with a genomic position (chromosome, position). When evaluating a variant, the variant’s genomic position is compared to the mapping table and the relevant publications are associated. If the variant has more than one publication, then it is reported as a moderate PS3.

#### PS4

The final strong pathogenic classifier looks at genome-wide association studies to identify variants with an odds ratio (OR) indicative of genotype-phenotype relationships. We use the UCSC gwasCatalog.txt file to generate a mapping table of chromosomes to position, and position to observed GWAS score (Hindorff et al., 2009).

To evaluate a variant, we check if the chromosome and position are in the mapping table and extract the value if it’s present. We weight the score from one to three depending on the OR and p-values, where a higher OR and lower p-value give the variant a higher score.

#### PM1

The first moderate pathogenic classifier evaluates if variants are within mutational hotspots and well-established functional domains. To evaluate domains that are mutational hotspots, we begin with the ClinVar variant VCF file. We start by building a mapping table associating domains with all variants seen in those domains. We then assign a score to each variant using the weights associated with the ClinVar review status. For each domain, we calculate the sum of all pathogenic variant scores, the sum of all benign variant scores, and the total of all scores. Using these values, we determine the pathogenic percentage and benign percentage for each domain. Domains with a pathogenic percentage greater than 80% and a total score of five (at least two variants) are considered mutational hotspot domains. These domains are stored using their genomic coordinates and, at runtime, are loaded into a mapping table.

If the variant is within one of these mutational hotspot domains, it meets the criteria and is given a score. The score is determined by the domain: default domains are scored supporting, domains with total score greater than 30 and pathogenic percentage greater than 90% are scored moderate, and domains with total score greater than 50 and pathogenic percentage greater than 95% are scored strong.

#### PM2

The second moderate pathogenic classifier looks at population database allele frequency to establish potential pathogenicity. For this, BIAS-2015 uses the gnomAD and 1000 Genomes data annotations from ICA. If a variant is missing from both gnomAD and 1000 Genomes, then the PM2 classifier is applied with a score of one. Or, if the variant was annotated with gnomAD or 1000 Genomes data and the allele frequency is below 1%, then the variant is assigned a supporting score.

#### PM4

This classifier evaluates in-frame insertions and deletions which, while not as detrimental as a null variant, are still indicative of pathogenicity. This is caveated by the qualifier that the variant should not be in a repeat region since in-frame indels in repeat regions are less likely to be pathogenic. We began by identifying repeat regions from the UCSC RepeatMasker track that intersect with the UCSC consensus coding regions (Kent, 2002; Pruitt et al., 2014; **?**; Smit et al., 2010). This is stored as a data file and loaded at runtime into a list of repeat regions.

We use the ICA consequences of ‘stop lost’, ‘inframe insertion’ and ‘inframe deletion’ to consider variants. If a variant has one of these consequences and is not in a repeat region, then it is assigned a base score of one. The variant is weighted based on the length of the indel: indels shorter than four amino acids are scored supporting, those between four and ten amino acids are moderate, and those which are greater than ten amino acids are strong.

#### PM5

The fifth moderate pathogenic classifier mirrors the first strong pathogenic classifier that compares amino acid changes from pathogenic variants. We use the same data extraction technique as in PS1 to define a list of pathogenic amino acids.

To classify a variant, we check if the gene and base amino acid is seen in our list of pathogenic gene/amino acid relations. If there is a match, the variant meets criteria and a score is assigned. The score can be incremented through two avenues that represent increased evidence: if the variant was reported as “pathogenic” or if there is sufficient weight associated with the ClinVar review status.

#### PP2

The second supporting pathogenic classifier evaluates if a missense variant is in a gene where missense variants are often associated with pathogenicity. To identify genes that are associated with missense pathogenicity, we again turned to ClinVar with a similar approach as in PM1. We began by creating a mapping table between genes and the variants associated with them. We then identified genes where over 80% of pathogenic variants are missense and, when looking at all missense variants in the gene, benign variants make up less than 10%. We only considered reviewed variants with an assigned ClinVar consequence. This list of genes is then saved as a text file which is loaded at runtime into a list.

To evaluate a variant for PP2, we have two conditions: first we check if the ICA-assigned gene name is in our list of qualifying genes; second, we check if the term ‘missense’ is in the ICA-assigned consequence. If both conditions are met, then the variant is assigned a supporting score.

#### PP3

This classifier is assigned based on computational pathogenicity predictive algorithms. For our implementation, we currently use three algorithms which are annotated by ICA: REVEL, DANN, and GERP (Ioannidis et al., 2016; Quang et al., 2015; Davydov et al., 2010). If a variant is found to have a pathogenic score in any of the algorithms, then it meets the criteria and is given a score.

While the 2015 ACMG standards specify this classifier can only be applied one time, subsequent SVI publications address it specifically emphasizing that in some cases these algorithms can have a strong predictive strength. To reflect this, we weight the PP3 classifier based on how many of the predictive algorithms concur on pathogenicity. A variant that meets cutoff levels for one term is scored supporting, for two terms it is scored moderate, and for all three it is scored strong.

Cutoff levels were derived from the respective algorithm publications. The authors are aware that subsequent work has been done to fine-tune these cutoff values for genomic applications, in some instances for PP3 specifically (Pejaver et al., 2022). We plan to incorporate these community discoveries in future versions of BIAS-2015.

#### PP5

ACMG recommends including classification data from multiple sources and using it as the basis for the PP5 classifier (Biesecker et al., 2018). For this classifier, we use a different schema than the other classifiers to incorporate the fact that some variants have an abundance of evidence that should not be overruled (Biesecker et al., 2018). Here we classify variants with stand-alone PP5 evidence if they have ClinVar review status ‘Practice Guidelines’ and ‘Reviewed by Expert Panel’. For remaining ClinVar variants, we reason that the evidence for their classification is not enough to overrule other lines of evidence. In these situations, we provide a PP5 score consistent with our other classifiers: weighing based on evidence. Variants with a review status of ‘criteria provided, conflicting interpretations’ are assigned a score of supporting, variants with a review status of ‘criteria provided, single submitter’ are assigned a score of moderate, and for variants with a review status of ‘criteria provided, multiple submitters, no conflicts’ are assigned a score of strong. Variants with any other review statuses were not viewed as credible and are not considered.

### Unimplemented Pathogenic Classifiers

BIAS-2015 does not directly evaluate PS2, PM3, PM6, PP1, and PP4. Users can provide information for each of these classifiers using the external call file.

Benign Classifiers

In the following sections, we describe how BIAS-2015 applies the ACMG criteria to each benign classifier. Figure 3 summarizes which benign classifiers are covered by BIAS-2015 and which should be handled using the optional external call file.

**Fig. 3.**
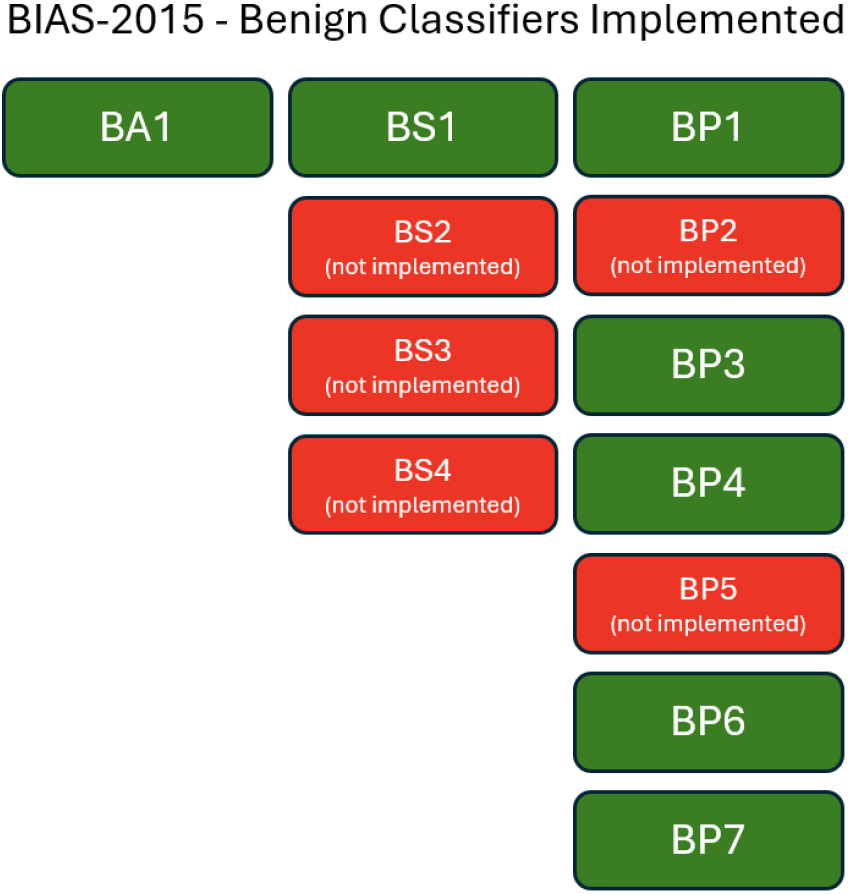
BIAS-2015 Benign Classifiers Implemented. The diagram shows the implementation status of various benign classifiers. Green boxes represent implemented classifiers, while red boxes with “not implemented” indicate classifiers that are not implemented.

#### BA1

This stand-alone classifier looks at population database allele frequency to establish penetrance of the variant across populations. The ACMG guidelines establish that variants with high penetrance across populations are unlikely to be pathogenic. ICA annotates variants with the gnomAD and 1000 Genomes frequencies. For BA1, a variant is applied the classifier if it has an allele frequency greater than 5% in either gnomAD or 1000 Genomes.

#### BS1

The first strong benign classifier applies the same population allele frequency annotation as the previous classifier, however, BS1 considers an allele frequency of 1%. If the variant frequency exceeds 1%, it is given a supporting score. This classifier can also be downgraded from strong to moderate and/or supporting in some scenarios when other conflicting evidence is present.

#### BP1

The first supporting benign classifier looks for missense variants within genes where only LOF mutations are associated with disease. We used the same data extraction technique as in PP2 to define a list of genes that are LOF only genes. If a variant’s gene name is in our list of LOF-only genes, and the ICA-assigned consequence is ‘missense’, then the variant is assigned a score of one.

#### BP3

This classifier excludes in-frame insertions and deletions in repeat regions from consideration, as these regions are known to be less pathogenic. We use the same data extraction technique as in PM4 to identify and exclude variants in these regions. If a variant has a consequence of ‘stop lost’, ‘inframe insertion’ or ‘inframe deletion’ and overlaps a repeat region, then the variant is assigned a score of one.

#### BP4

This classifier is used to rule out benign variants that are non-functional and predicted benign by *in silico* predictive algorithms. We use the same data extraction technique as in PP3 to generate a mapping table from *in silico* predictive algorithms. For BP4, however, we consider a low probability score for all algorithms. Variants that meet the cutoff levels for benign (set forth by the algorithm publications) for all three predictive algorithms are assigned a strong score, variants meeting benign levels for two algorithms are assigned a moderate score, and variants meeting benign levels for one algorithm are assigned a supporting score.

#### BP6

This classifier rules out benign variants based on established clinical data. For this classifier, we use the same data extraction technique as in PP5 to generate a mapping table of ClinVar reviews. We follow the same logic to establish ClinVar classification levels.

#### BP7

This classifier rules out benign synonymous variants. For this classifier, we rely on ICA annotations and apply a score of one for synonymous variants. If a variant is synonymous and has a low probability score in all three predictive algorithms, it is assigned a score of three.

### Unimplemented Benign Classifiers

BIAS-2015 does not directly evaluate BP2, BP5, BS2, BS3, and BS4. Users can provide information for each of these classifiers using the external call file.

## Results

Consider variant NM 000257.3(MYH7):c.1207C>T with genomic coordinate chr14, 23898488, G>A on build hg19 as it is processed by the BIAS-2015 algorithm. This missense variant changes the amino acid from an arginine to a tryptophan, as noted by the ICA annotation. When processing this variant with BIAS-2015, the algorithm applied the following classifiers: PS1, PM1, PM2, PP2, PP3, PP5, and BP4. Additionally, each classifier is justified as demonstrated in Table 1.

**Table 1.** Classifier Scores and Justifications. This table provides the scores assigned to various classifiers based on specific justifications. Each classifier (PS1, PM1, PM2, PP2, PP3, PP5, BP4) is evaluated with a score and the reasoning behind the score, including amino acid changes, domain-specific information, population data, variant consequences, computational evidence, and clinical reports.

| Code | Score | Justification |
| --- | --- | --- |
| PS1 | 3 | Amino acid change R403W in gene MYH7 is associated with ClinVar Pathogenic variant rs3218714 with review status 'reviewed by expert panel'. |
| PM1 | 2 | Variant is in domain, P12883 where 0.9067% variants are associated with pathogenicity (var score 175). |
| PM2 | 1 | Variant is missing from gnomAD. |
| PP2 | 1 | Missense variant consequence missense_variant in MYH7 which was found to have a low rate of missense variation and where missense variants are a common mechanism of disease. |
| PP3 | 1 | Computational evidence supports a deleterious effect; GERP -0.787 — DANN 1 — REVEL 0.822. |
| PP5 | 5 | Variant was found reported as pathogenic in ClinVar with review status of 'reviewed by expert panel'. |
| BP4 | 1 | Computational evidence supports a benign effect; GERP -0.787 — DANN 1 — REVEL 0.822. |

We recognize that this variant is reported in ClinVar as pathogenic with a review status of ‘Reviewed by expert panel’, and the algorithm has recognized this and weighted it with a stand-alone value, ensuring this variant will be classified in P/LP. Additionally, we note that BIAS-2015 would classify this variant as pathogenic even if the PP5 classifier was excluded.

### Analysis of eRepo Data

ClinGen’s Evidence Repository (eRepo) serves as a pivotal resource for the clinical interpretation of genetic variants, offering meticulously curated evidence regarding their clinical significance. Recognized as a robust and reliable source of truth for ACMG classifications, eRepo stands out due to its rigorous curation process endorsed by the FDA. Variants included in eRepo are reviewed and annotated using the ClinGen Variant Curation Interface (VCI), which adheres to the standardized ACMG/AMP variant classification guidelines. Each variant undergoes an expert review where evidence criteria are manually applied, ensuring a comprehensive and accurate classification. The VCI facilitates collaboration among ClinGen Expert Panels, enabling them to evaluate and incorporate relevant clinical, genetic, population, and functional evidence. This structured process not only supports the clinical validity of the classifications but also ensures transparency and consistency. The finalized variant classifications are then published in the eRepo and ClinVar databases, allowing for broad accessibility and peer review within the genomics community.

Crucially, the eRepo database can be downloaded as a TSV file, containing detailed variant information, including manually applied ACMG classifiers. We began our validation by generating an hg19 VCF file from the eRepo database; 7,324 out of 7,632 eRepo entries were successfully converted to VCF format. We then took this VCF file and classified all variants in it using BIAS-2015. When we compared the classification of the BIAS-2015 algorithm with the classifications provided in eRepo, we were able to perform this comparison for 7,110 out of 7,324 variants. This was done in a completely automated manner, and all code and commands executed to perform this comparison have been provided in our GitHub repository along with all data files used and generated during this process are available on our GitHub data repository (Bitscopic, 2024).

To establish the broad performance of BIAS-2015, we calculated values for sensitivity and specificity across three categories: benign/likely benign (B/LB), variants of uncertain significance (VUS), and pathogenic/likely pathogenic (P/LP). The scores are reported in Table 2.

**Table 2.**
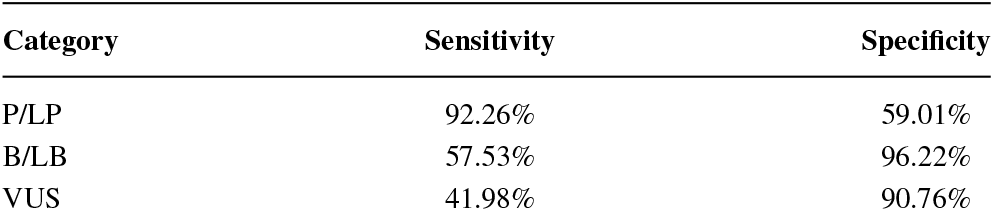
Sensitivity and Specificity of Different Categories. This table presents the sensitivity and specificity percentages for three categories: Pathogenic/Likely Pathogenic (P/LP), Benign/Likely Benign (B/LB), and Variants of Uncertain Significance (VUS). The sensitivity and specificity values indicate the accuracy of classification for each category.

While most ACMG classifier algorithm comparisons end here, both BIAS-2015 and eRepo use the same ACMG classifier codes and both provide written explanations for every classifier; therefore, we can evaluate each ACMG classifier to determine whether BIAS-2015 is in concordance with the eRepo database. To explore this, we evaluated the number of classifiers assigned by both algorithms when using the eRepo generated VCF file with 7,324 entries and compared the number of assignments for each classifier in Figure 4.

**Fig. 4.**
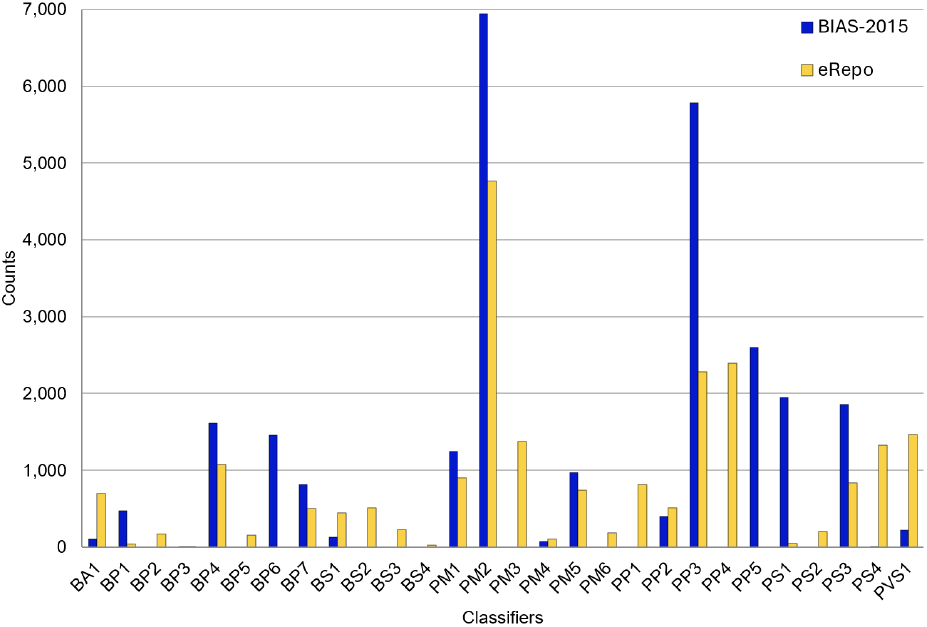
Counts of ACMG Criteria by BIAS-2015 and eRepo. This bar chart compares the counts of various ACMG criteria classifiers between BIAS-2015 (blue) and eRepo (yellow). The x-axis lists the classifiers, while the y-axis indicates the counts.

Our initial analysis revealed that in some classifiers there is concordance, whereas in others, there is a large disparity. Since BIAS-2015 does not evaluate BP2, BP5, BS2, BS3, BS4, PM3, PM6, PP1, PP4, and PS2, the eRepo dataset will always be discordant on these classifiers. Additionally, eRepo does not assign BP6 and PP5 classifiers to any variants, so BIAS-2015 will never be concordant on these classifiers. However, even within classifiers that both BIAS-2015 and eRepo evaluate, there are widespread differences. For instance, when comparing PVS1 classifier calls, BIAS-2015 assigns PVS1 to 221 variants, yet eRepo assigns it to 1,476 variants. This highlights the need for refinement in the PVS1 classifier, specifically in the inclusion criteria handled by the preprocessing generation of a LOF gene list. Another example is in PP3, where BIAS-2015 classifies variants 5,787 times and the eRepo only assigns it 2,283 times. This indicates that BIAS-2015 is being too lenient in its computational pathogenicity algorithm cutoffs and more stringent values should be used. These insights highlight that further improvements are needed to refine the automated classification process.

## Discussion

### Manual Adjustment

While BIAS-2015 can perform an automated analysis, the authors stress that for the most accurate classifications, a user should manually provide classifiers for the codes BIAS-2015 does not evaluate. Additionally, users can review the weights that BIAS-2015 applies to every classifier and adjust them if appropriate. This can be done using the optional external call file when running the algorithm.

The optional external call file is the same format as the BIAS-2015 output, meaning a user can include their own scores or classifications with BIAS-2015 in a three-step fashion:

1. The user runs BIAS-2015 on their default VCF file to generate a BIAS-2015 output.
2. The user copies this output file and manually/programmatically inserts/adjusts any applicable ACMG codes using their own expertise. The user saves the modified file as an external call file.
3. The user runs BIAS-2015 a second time, this time providing their external call file as an argument. BIAS-2015 will recalculate the variant codes, load the user-provided codes and weights, and then classify the variants through the BIAS-2015 algorithm.

### Algorithmic Refinement

Currently there are many algorithmic approaches to variant classification that vary from AI-modeling to attempting to fit to a continuous latent scale of benign-pathogenic. While these algorithms are impressive in their performance, they do not report the individual ACMG codes and in turn do not provide clear justification for each code.

Our motive to create our tool in this manner was driven in part by new community databases such as eRepo, which annotate all variants with detailed ACMG code information. Since BIAS-2015 also annotates all variants with detailed ACMG code information, we can compare the codes BIAS-2015 assigns to those the eRepo assigns and perpetually fine-tune the algorithm.

In Table 3 we compare BIAS-2015 on the same example variant from before, NM 000257.3(MYH7):c.1207C>T, this time showing the eRepo classifier codes and their justification side by side with the BIAS-2015 classifier codes.

**Table 3.** Detailed Comparison of eRepo and Bitscopic Classifiers with Corresponding Justifications for Pathogenicity Assessment. The table compares specific criteria codes and justifications used in two systems: eRepo and Bitscopic. For each classifier (e.g., PS1, PM1), the table outlines the evidence used by eRepo and Bitscopic, highlighting differences in the interpretation and sources of pathogenicity evidence.

| eRepo Code | eRepo Justification | Bitscopic Code | Bitscopic Justification |
| --- | --- | --- | --- |
| PS1 | Amino acid change R403W in gene MYH7 is associated with ClinVar pathogenic variant. | PS1 | Variant is in domain, P12883, where 0.9067% variants are associated with pathogenicity. |
| PM1 | This variant lies in the head region of the protein (aa 181-937) and missense variants in this region are statistically more likely to be disease-associated. | PM1 | Variant is in domain, P12883, where 0.9067% variants are associated with pathogenicity. |
| PM2 | Variant was absent from large population studies (PM2; http://exac.broadinstitute.org). | PM2 | Variant is missing from gnomAD. |
| PP3 | Computational prediction tools and conservation analysis suggest that this variant may impact the protein. | PP3 | Computational evidence supports a deleterious effect. |

In this example, we see a degree of concordance between BIAS-2015 and the expert-assigned codes; however, there are notable differences. BIAS-2015 applies the PS1 code to this variant, whereas the eRepo experts use the PM5 code. Both codes look at the amino acid change associated with a variant and ask if the amino acid has been associated with known variants. While BIAS did not assign the correct code, it did correctly note that the amino acid change is known to be pathogenic.

Another deviation noted by the authors is that BIAS-2015 includes an unwarranted benign classifier, BP4. Upon investigation, this is explained due to the cutoff values used in the PP3 and BP4 implementations which were recommended in their base publications. This brings to our attention the need to re-evaluate the PP3 and BP4 algorithmic implementations and further refine them.

### Performance Interpretation

When evaluating the eRepo dataset, BIAS-2015 achieved a benign specificity of 96.22%. This can be interpreted as out of 1,000 variants that are not benign, we call 962 of these variants not benign. Inverting these numbers, BIAS is incorrectly calling 38 variants benign out of every 1,000 benign variants. We attribute this high specificity to the order of the ACMG combining criteria and the inclusion of our BIAS-2015 conflicting classifications VUS combining criteria.

The authors also recognize that there is extreme interest in the bioinformatics research and medical community surrounding detection of *de novo* pathogenic variants. When evaluating this, BIAS-2015 was able to correctly identify 92.26% of pathogenic variants in the eRepo dataset. The authors noted that this number includes variants that are reported in ClinVar and will therefore get classified with our PP5 code. To evaluate BIAS-2015 performance on variants that are not directly in ClinVar (similar to *de novo* variants), we turned off the PP5 classifier and reran our analysis against the eRepo dataset. In this scenario, BIAS-2015 reported a pathogenic sensitivity of 81.80%.

BIAS-2015 performs poorly in benign variant sensitivity (57.53%) and in pathogenic specificity (59.01%). We again attribute this to the order in which BIAS-2015 evaluates the ACMG combining criteria and the inclusion of the BIAS-2015 conflicting classification VUS combining criteria. These numbers highlight the need for further algorithmic refinement.

## Data Availability

https://github.com/Bitscopic/BIAS-2015

https://erepo.clinicalgenome.org/evrepo/

## Declaration of Interests

The authors declare the following conflict of interest: The BIAS-2015 algorithm described in this paper is intended for use in a commercial product by Bitscopic. This potential financial interest does not alter the authors’ adherence to community best practices and scientific standards in developing and validating the algorithm. The development of BIAS-2015 was conducted independently by Bitscopic, and no external funding or influence was received from commercial entities outside of this potential interest.

## Acknowledgements

Special thanks to Illumina for building and providing a robust annotation tool, Illumina Connected Annotations, which was instrumental in the development of this algorithm. We would also like to acknowledge the UCSC Genome Browser team for their comprehensive and accessible genomic data resources, which were critical to the preprocessing phase of our algorithm.

## Author Contributions

- CE: Conceptualization, Methodology, Software, Validation, Writing, Review & Editing.
- JM: Review & Editing, Visualization, Writing.
- RB: Review & Editing, Visualization, Writing.
- VB: Review & Editing, Writing.

## Data and Code Availability

The algorithm code, preprocessing code, and validation code are all made available on GitHub, promoting transparency and accessibility for the broader scientific community (Bitscopic, 2024).

## Notes

### Competing Interest Statement

The authors have declared no competing interest.

### Funding Statement

Bitscopic internal funding.

